# Estimation of cancer cases in transgender and gender diverse people in England

**DOI:** 10.64898/2026.04.21.26351378

**Authors:** Chloé Pasin, Sarah S Jackson, Lilly-Emma Thynne, Brigit McWade, Tanith Westerman, Rue Ball, James Kavanagh, Stewart O’Callaghan, Kyle Ring, Chloe Orkin, Alison M Berner

**Affiliations:** SHARE Collaborative, Blizard Institute, Queen Mary University of London, London, UK; Lancaster University, Lancaster, UK; Barts Cancer Institute, Queen Mary University of London, London, UK; Synnovis Analytics, London, UK; Wolfson Institute of Population Health, Queen Mary University of London, London, UK; OUTpatients Charity, London, UK; Barts Health NHS Trust, London, UK; Chelsea & Westminster Hospitals NHS Trust, London, UK

## Abstract

**Objectives:** To estimate current, and 5- and 10-year projected, number of cases of cancer per year in transgender and gender diverse (TGD) people in England, overall and by tumour type, accounting for uptake of gender affirming care (GAC).

**Design:** Population-based epidemiological modelling study using an age-stratified Monte Carlo simulations approach and the NORDPRED method for predictions.

**Setting:** Models estimating cancer case numbers for TGD people in England based on publicly available 2023 cancer surveillance data and survey-based 2025 GAC access, and predicted at 5 and 10 years hence.

**Participants:** TGD people aged 15 years and above.

**Main outcome measures:** Primary cancer cases per year overall, by gender, age group, tumour type, and current and planned GAC.

**Results:** The estimated TGD population size in England is 441547 (95% uncertainty interval (UI) 429207- 452890). Total cases per year of cancer in TGD people is expected to be 966 (95% UI 882-1069) excluding non-melanoma skin. Most cases are expected to occur in people aged 60-64. The top 5 expected cancers in TGD people are breast (19%, n = 187, 95% UI 149-241), colorectal (12%, n = 117, 95% UI 106-129), lung (11%, n = 108, 95% UI 96-122), melanoma (7.1%, n = 69, 95% UI 64-74) and urinary (6.2%, n = 60, 95% UI 54-67). Total cases of cancer in TGD people are estimated to be 1740 (95% UI 1584-1934) in 5 years and 2258 (95% UI 2066-2507) in 10 years (excluding non-melanoma skin). If TGD people were able to access their planned level of GAC, this would reduce these figures to 1555 (95% CI 1386-1766) and 2012 (95% CI 1797-2282) respectively.

**Conclusions:** This study provides prediction of cancer cases in TGD people in England, supporting the planning of service provision and training. This is vital, as with increasing disclosure, and long wait times for GAC, cancer cases in TGD people are predicted to increase.

**Summary Boxes:** *What is already known on this topic:* The annual number of cases of cancer in transgender and gender diverse (TGD) people in England is currently unknown as gender incongruence is not collected as part of the National Cancer Registration and Analysis Service. Some gender-affirming care (GAC) interventions are known to modulate cancer risk. Use of testosterone and chest reconstruction for transmasculine people is known to reduce their incidence of breast cancer compared to cisgender women. Use of oestradiol alongside medical or surgical androgen suppression has been shown to reduce the incidence of prostate cancer in transfeminine people while increasing their risk of breast cancer, compared to cisgender men.

*What this study adds:* This study found that there are likely to be approximately 966 cases of cancer (excluding non-melanoma skin) in TGD people per year in the UK. Though total annual cases of cancer in TGD people are expected to be 2258 in 10 years, improved access to gender-affirming care could reduce total cases to 2012 (a 11% reduction). These figures provide additional justification for funding to improve access to GAC via the National Health Service (NHS), as well as for training on the oncological needs of this population.

## Background

An estimated 0.5-0.87% of individuals in the United Kingdom (UK)[1,2] are transgender or gender diverse (TGD) (Table 1). The largest proportion of TGD individuals are reported in younger age groups, with 2.4% of people aged 15-19 years identifying as TGD, suggesting increasing disclosure in successive generations.[1] Though gender is recorded and reported by the National Cancer Registration and Analysis Service (NCRAS), it is not recorded separately from sex assigned at birth as the terms ‘sex’ and ‘gender’ are frequently conflated in electronic health records in the UK. Gender incongruence (Table 1) is also not recorded. We therefore lack robust cancer incidence data for TGD people.

**Table 1:** Definitions relevant to transgender and gender diverse (TGD) populations and their health.

| **Term** | **Definitions** |
| --- | --- |
| Cisgender | The term cisgender describes an individual whose lived experiences of gender align with traditional expectations associated with their sex assigned at birth.[3] |
| Gender | Gender can refer to a group of people who share qualities, characteristics or behavioural traits that society associates with being female, male or gender diverse.[4] |
| Gender Affirming Care (GAC) | The term gender affirming care, also known as gender affirming healthcare and transgender healthcare, refers to inter- and multidisciplinary care between endocrinology, surgery, voice and communication, primary care, reproductive health, sexual health and mental health interventions that support the individual to affirm their gender experiences. Not all TGD people will seek healthcare interventions.[5] In this paper, we refer to hormonal and surgical GAC. |
| Gender Dysphoria | The term gender dysphoria describes experiences of psychological and physiological discomfort or distress associated with a discrepancy between an individual's gender and their sex assigned at birth.[6] |
| Gender Incongruence | The revised World Health Organisation International Classification of Diseases (ICD-11) defines gender incongruence as “a marked and persistent incongruence between an individual's experienced gender and the assigned sex”.[7] |
| Non-binary (person) | A phrase used refer to those with gender identities outside the gender binary.[5] |
| Transgender and Gender Diverse | The phrase used to describe a diversity of lived experiences among communities of people whose gender is different to the gender socially attributed to their sex assigned at birth.[5] |
| Transfeminine (person) | A phrase used to describe someone whose gender is more female than male and was assigned male at birth. |
| Transgender man | A man who was assigned female at birth. |
| Transgender woman | A woman who was assigned male at birth. |
| Transmasculine (person) | A phrase used to describe someone whose gender is more male than female and was assigned female at birth. |

Cancer incidence is likely to differ in TGD people compared to cisgender people (Table 1) for several reasons. Firstly, evidence from the UK Clinical Practice Research Datalink (CPRD) suggests that TGD people have a higher prevalence of cancer risk factors including obesity, alcohol use, smoking and blood-borne virus infection.[8] Secondly, gender-affirming hormone therapies and surgeries may modulate risks of sex-specific cancers. Observational studies from the Netherlands indicate that the risk of breast cancer in transgender men is elevated compared to cisgender men, but lower than that seen in cisgender women,[9] owing to testosterone therapy and removal of a large proportion of breast tissue at masculinising chest surgery. In transgender women, the risk of breast cancer is lower than that of cisgender women but elevated compared to cisgender men, owing to increased circulating oestradiol and breast tissue development.[9] Though the prostate is not removed as part of genital reconstructive surgery, transgender women taking oestradiol and androgen deprivation therapy, with or without orchiectomy, have a reduced prostate cancer risk compared to cisgender men.[10] Currently, there is no evidence of an increased risk for gynaecological cancers owing to gender-affirming hormones.[11,12] However, However, transgender men who undergo hysterectomy and bilateral salpingoophorectomy with or without genital reconstruction reduce the risk of gynaecological cancers to almost zero.[5] (Note that in rare cases ovarian or primary peritoneal cancers may only become clinically evident years after bilateral salpingoophorectomy). [11,12]

Thirdly, long NHS waiting times delay access to gender-affirming care (GAC) for many TGD people in the UK. [13,14] Because of these delays, many TGD individuals instead self-fund care privately in the UK, by travelling abroad, or self-medicate with hormones from unregulated sources without medical oversight.[13,15] Estimating current or previous GAC from health records can be difficult, affecting incidence estimates for many cancers. Lastly, high levels of anticipated and experienced stigma and discrimination can lead to healthcare mistrust,[16,17] impacting engagement with screening and preventative care.[18,19] Gender dysphoria (Table 1) can reduce engagement with sex-specific screening programmes (e.g. breast, and cervical cancer), and highly-gendered information materials and clinic environments.[18,20,21] Together, these factors make it challenging to estimate the cancer incidence among TGD people, and to plan appropriate service provision and training for the oncology workforce. We set out to estimate the annual number of cancer cases in TGD people in the UK, overall and by tumour type, using publicly available data and to model cancer cases over the next 5 and 10 years.

## Methods

### Data sources

For all data sources, we used the most recent available. Age and sex-specific cancer incidence with corresponding confidence intervals in the cisgender population were obtained from the 2023 National Disease Registration Service cancer registry.[22] For ease, primary malignant cancer types were grouped by anatomical site (see Supplementary Table 1). Benign and secondary cancers were excluded.

Age- and sex-specific denominators for cisgender people were obtained from the Census 2021 counts through the Office of National Statistics (ONS).[2] To obtain the denominator for the age-specific proportion of TGD people in England, we used the NHS GP Patient Survey (GPPS) 2025.[1] Please see supplementary methods for further details about these datasets.

Given the lack of publicly available data on access to gender-affirming care, we relied on an online mixed-methods community survey with 419 respondents, as this also captures those who may source this care outside of the NHS (private providers and self-medication).[23] Gender-affirming interventions that may alter cancer risk, alongside percentage of people currently accessing and planning to access these, and median age of access, is shown in Supplementary Table 2. Due to long waiting lists for GAC in the NHS, there were large discrepancies in gender-affirming care people were accessing currently and those that they wished to access in the future (Supplementary Table 2).

Post-GAC standardised-incidence ratios were obtained from the literature.[9,10] Supplementary Table 3 summarises the data and the corresponding references. [24]

We estimated the expected number of cancer cases in TGD populations in England using an age-stratified Monte Carlo simulations approach. For each cancer site and patient group, we combined:

i. Age-specific cancer incidence in the reference cisgender population (defined as male or female);
ii. Age-specific population counts of TGD people, obtained by combining age-specific population counts of male and female in England and the age-specific proportion of TGD people in the population;
iii. Estimation of access and planned access to gender-affirming hormones and surgery in TGD people;
iv. Estimated post-GAC standardised-incidence ratio (SIR).

For each parameter, we sampled from a pre-defined parametric distribution. Results were summarised by age category and in total.

### Estimation of cancer cases

For each age stratum labelled j=1, …, J (j=1: age 15 to 19, j=2: age 20 to 24, …, j=J: age 90 and more), we estimated the number of cancer cases using the following formula:

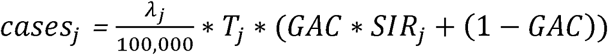

With λ*_j_* the cancer incidence in the reference cisgender population, *T_j_* the number of TGD people, GAC the GAC rate (not age-dependent, for hormonal and surgical GAC affecting cancer risk) and S/R_j_ the standardized-incidence ratio.

When SIR > 0, *SIR_j_* is set at the corresponding value and is constant across age strata. In the case of SIR = 0, we considered an age-dependent SIR, which *SIR_j_* =1 until median age of surgery, and *SIR_j_* =0 after. For cancers where GAC does not affect cancer risk, *SUR* and *SIR_j_* were set at 1.

### Parametric distributions

Uncertainty was considered by independently sampling each parameter using the following distributions:

- Cancer incidence λ*_j_* was drawn using a log-normal distribution with mean value = log(rate) and standard deviation = (log(UCI)-log(LCI))/(2*1.96), with UCI and LCI the upper and lower reported confidence intervals. If the reported rate was = 0 with LCI = 0 and UCI > 0, we used a log-uniform distribution on the interval [10^-6^;UCI]. Cancer incidence in the cisgender female population was used as a reference for transgender men and non-binary people assigned female at birth. Cancer incidence in the cisgender male population was used as a reference for transgender women and non-binary people assigned male at birth.
- Prevalence of TGD people was drawn using a beta distribution *p_j_ ∼ Beta*(*k_j_* + 1, *n_j_* - *k_j_* + 1) with *k_j_* the number of transgender or non-binary people in the GPPS survey and n_j_ the total population in the GPPS survey in this age stratum. In the GPPS survey, numbers were provided per age strata of 10 years (while cancer incidence was provided per age strata of 5 years). To harmonise the age categories, we considered a uniform repartition in the 10-years age strata (i.e. we divided by two to obtain the population numbers in the two corresponding 5-years age strata).
- Age-specific population total counts were obtained by adding female and male counts obtained from the census and drawn from a normal distribution: *Nj = N_Fj_ + N_Mj_* with *N_Fj_*∼ *N*(*μ_Fj_, σ_Fj_*) and *N_Mj_*∼ *N*(*μ_Mj_, σ_Mj_*).
- TGD population counts were obtained by drawing from a binomial distribution *T_j_* ∼ *Binomial*(*N_j_, p_j_* ).
- Surgery rates (either accessed, or planned and accessed) were obtained by drawing from a beta distribution *SUR ∼ Beta*(*k_SUR_* + 1, *n_SUR_* - *k_SUR_* + 1) with *k_SUR_* the number of TGD accessing (or planning and accessing) gender-affirming surgeries and n_SUR_ the total population included in the study.
- For SIR>0, we used a log-normal distribution with mean value = log(SIR) and standard deviation = (log(UCI)-log(LCI))/(2*1.96), with UCI and LCI the upper and lower reported confidence intervals.

### Monte Carlo simulations

We ran 1,000 Monte Carlo simulations for each cancer, sub-population, GAC category (accessed, or accessed and planned i.e. simulating the desired level of access). Cancer cases were obtained for each age stratum and added within each simulation set to obtain the total number of cancer cases across all age strata and sub-populations. Total expected cases were obtained by computing the mean among all Monte Carlo simulations and empirical 95% uncertainty intervals were computed.

### 5- and 10-year predictions (2031, 2036)

We estimated the number of cancer cases in TGD populations in England by using the same Monte Carlo simulation method.

#### Data sources and parametric distributions

Cancer incidence in the time periods 2029-2033 and 2034-2038 were predicted using the NORDPRED method with a power-5 link function.[25] The method was implemented using source code from the Norwegian Institute of Public Health.[26] Age- and sex-specific projections were made using historical cancer incidence rates (from 2004-2023) aggregated in 5-year time periods, and ONS future population estimate in England from the principal census scenario, with population numbers aggregated over 5-year period matching the cancer incidence data structure. Incidence rates were sampled from log-normal distributions with NORDPRED predictions as mean value and considering a coefficient of variation of 7.5% (CV = 0.075), given previous work suggesting a 5-10% relative bias in predictions.[27]

Age- and sex-specific denominators for cisgender people were obtained from ONS future population estimate in England. No sampling uncertainty was applied to the age-specific population counts, but predictions were obtained under three scenarios: principal, low fertility, mortality and migration and high fertility, mortality and migration.

We shifted the prevalence of TGD populations by 5 and 10 years: numbers from the 2025 GPPS survey were re-assigned to the next higher age category (either +5 or +10 years). The youngest age category (15-19) was assumed to stay constant at the values from the 2025 GPPS survey. Prevalence of TGD people was similarly sampled from a beta distribution, and TGD counts from a binomial distribution.

We used current accessed surgery rate, accessed and planned rates and post-GAC SIR as previously.

#### Size of the TGD population

We estimated the number of TGD people in England by using the same Monte Carlo simulation method and drawing TGD population counts were obtained from a binomial distribution *T_j_* ∼ Binomial(*N_j_*, *p_j_* ) as previously indicated. Population counts were obtained for each age stratum and added within each simulation set to obtain the total number of people across all age strata and sub-populations. Total expected population counts were obtained by computing the mean among all Monte Carlo simulations and empirical 95% uncertainty intervals were computed.

#### Implementation and code

Analyses were conducted in R version 4.4.2 using parallel computing for the Monte Carlo simulations. Code and source datasets are available on GitHub for reproduction: https://github.com/chlpasin/Estimation-of-cancer-cases-in-TGD-people-in-England

## Results

We estimated a total of 441547 TGD people in England (UI 429207- 452980); 114803 trans women (UI 109434-120826), 161770 trans men (UI 154757-168326), and 164974 non-binary people (UI 154746-176352). A breakdown by age is shown in Supplementary Figure 1.

The total cases per year of cancer in TGD people was expected to be 966 (95% uncertainty interval (UI) 882-1069) excluding non-melanoma skin and 1542 (95% UI 1401-1707) including non-melanoma skin (Supplementary table 4). The age distribution of cases is shown in Figure 1A, with the most cases expected to occur in people aged 60-64 (166 cases, 95% UI 132-197, including non-melanoma skin) (Supplementary table 5).

**Figure 1.**
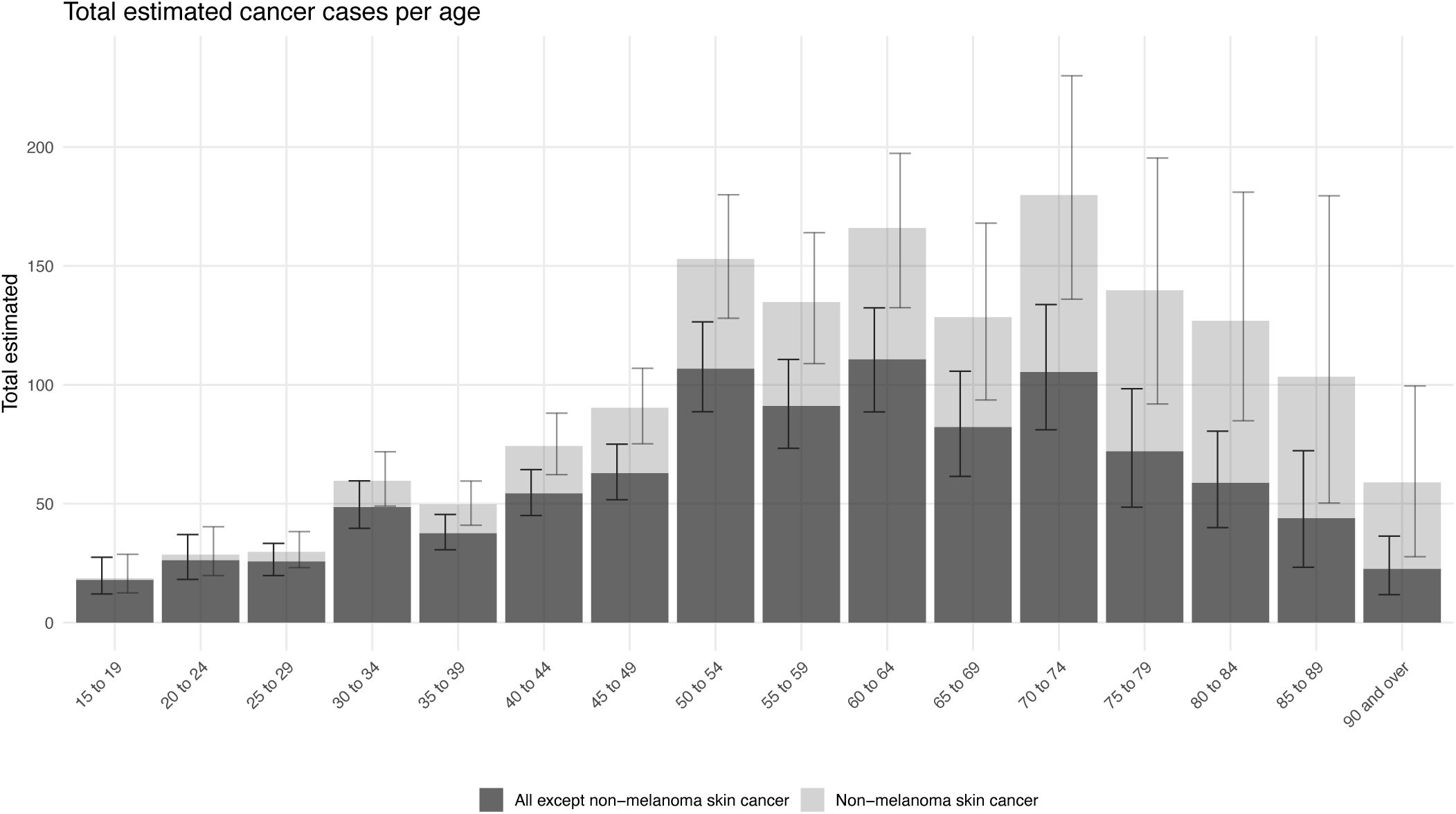
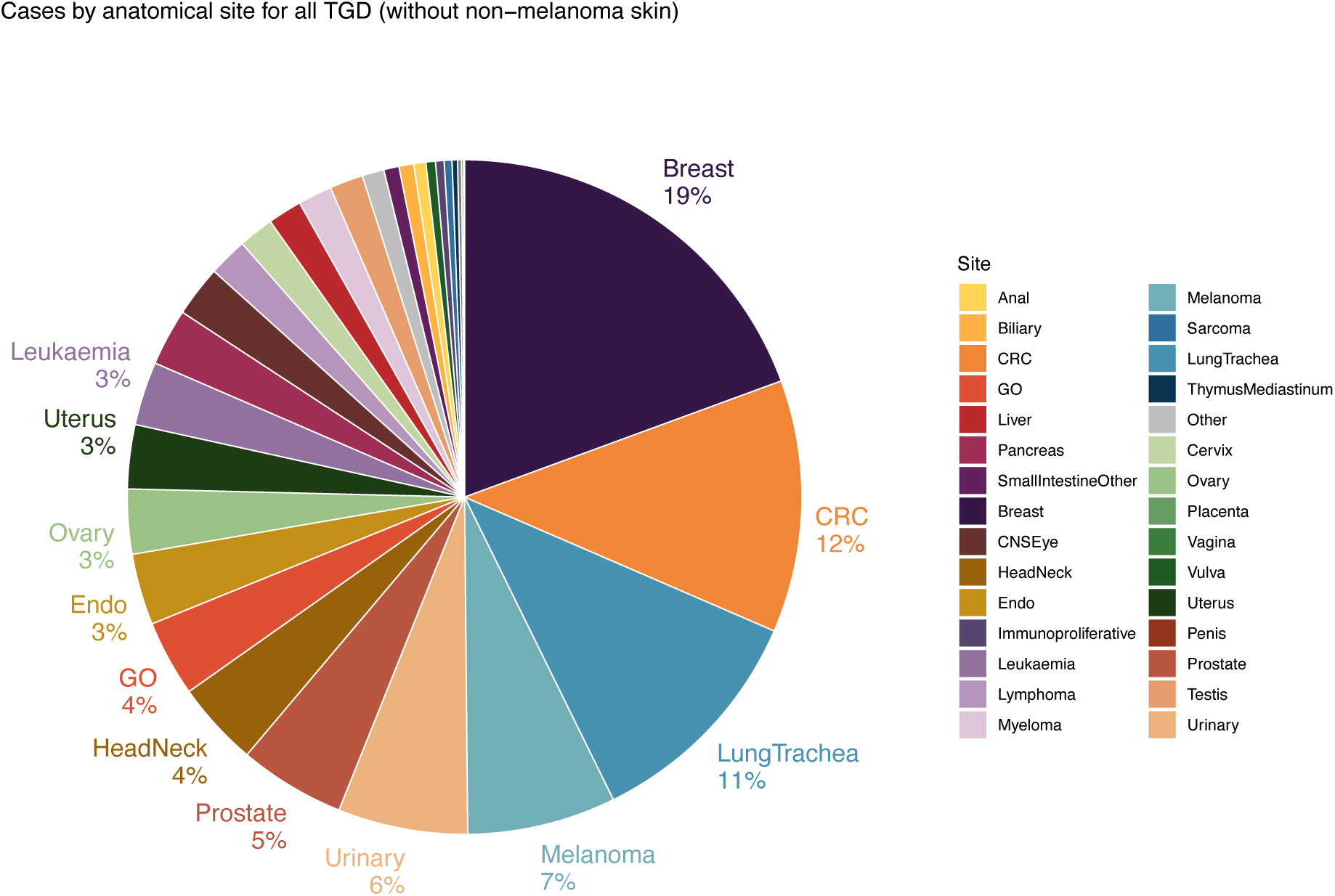
A. Histogram showing the estimated annual number of cancer cases in TGD people (y-axis). Each bar corresponds to an age category (x-axis). Error bars show 95% uncertainty intervals for each age category. Dark grey shows the number of cases across all cancer type, except non-melanoma skin cancer. Light grey shows the total number of cases (including non-melanoma skin cancer). B. Pie chart depicting the proportion of total estimated number of cancer cases for all TGD by anatomical site. Each colour corresponds to a different site, as indicated in the legend. Non-melanoma skin cancers are not included in this plot.

Excluding non-melanoma skin, the majority of cancer cases are expected to occur in transgender men (n = 423, 95% UI 389-461) followed by transgender women (n = 372, 95% UI 329-431), non-binary people who were assigned female at birth (n = 115, 95% UI 103-129) and non-binary people who were assigned male at birth (n = 56, 95% UI 46-69). Expected case estimates by age across these four groups are shown in Figure 2A-D. While cases in transgender men and transgender women occupy a distribution typical for the cisgender population with cancer cases peaking in older age groups (Figure 2A,C, Supplementary Figure 2A-C), this is not the case for non-binary populations (Figure 2B, D Supplementary Figure 1A-C).

**Figure 2.**
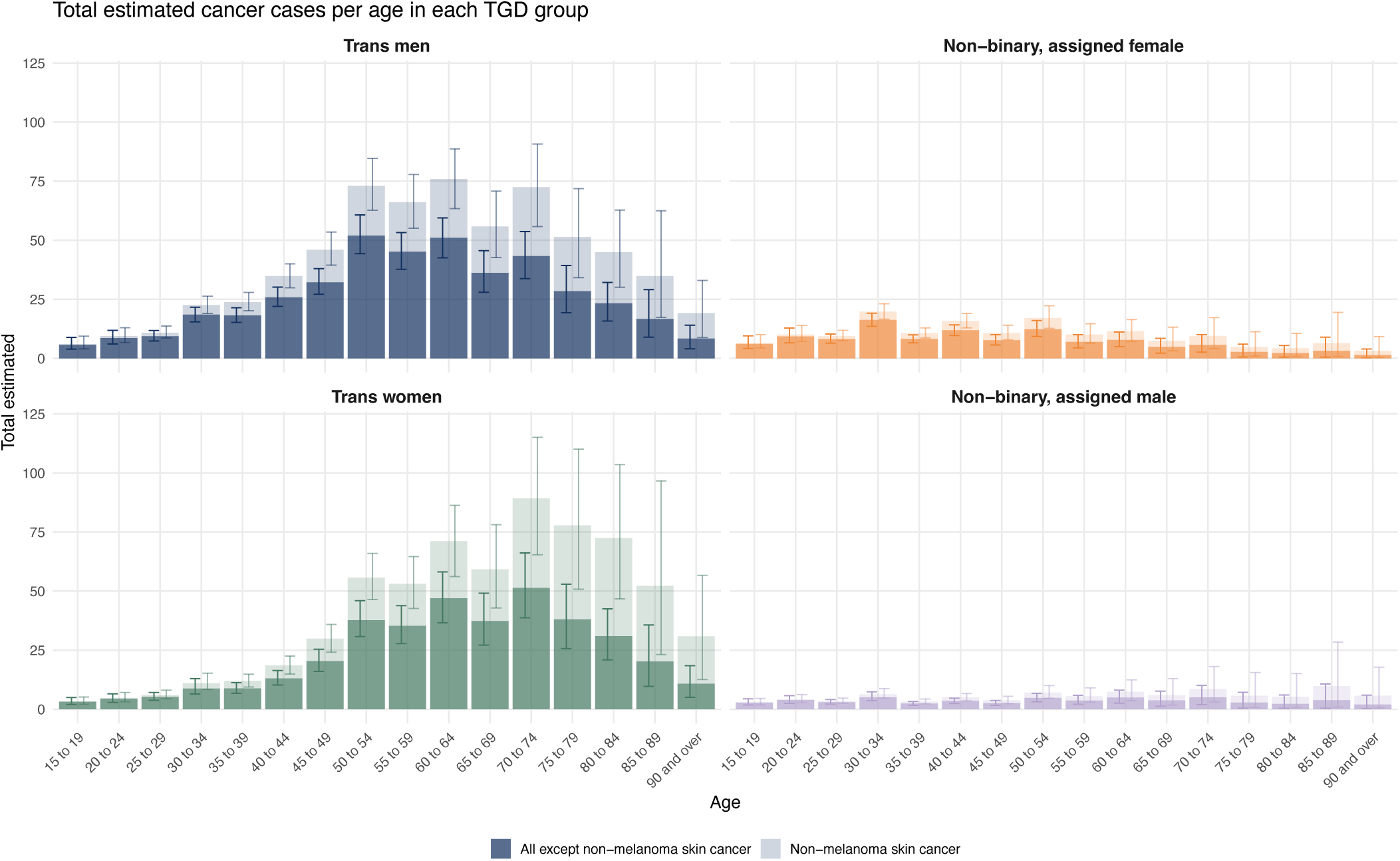
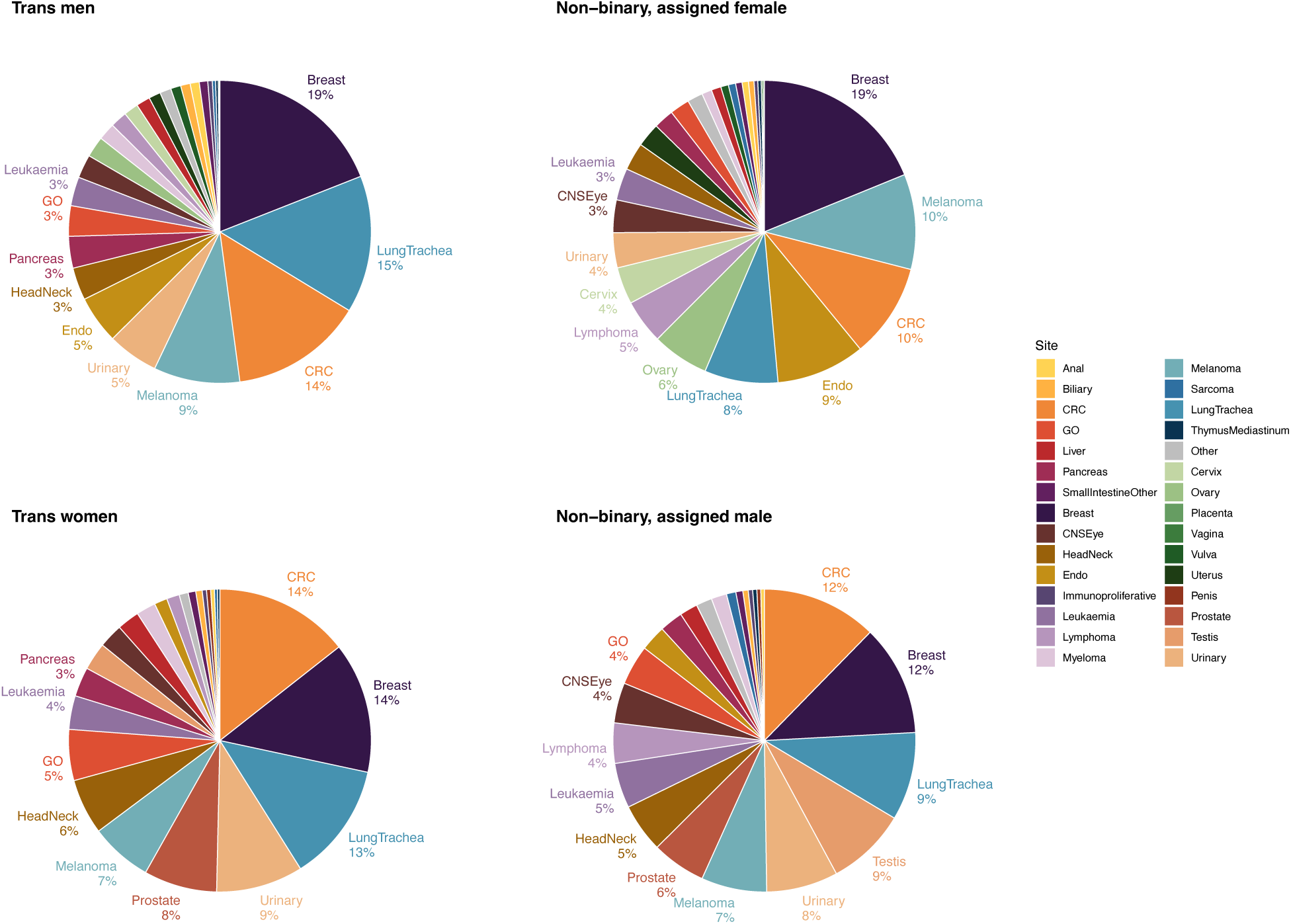
A-D. Histogram showing the estimated number of cancer cases in 2025 (y-axis) in transgender men (A, top left, dark blue), non-binary people assigned female at birth (B, top right, orange), transgender women (C, bottom left, dark green), and non-binary people assigned male at birth (D, bottom right, purple). Each bar corresponds to an age category (x-axis). Error bars show 95% uncertainty intervals for each age category. Dark colour shows the number of cases across all cancer type, except non-melanoma skin cancer. Light colour shows the total number of cases (including non-melanoma skin cancer). E-G. Pie chart depicting the proportion of total estimated number of cancer cases by anatomical site in transgender men (E, top left), non-binary people assigned female at birth (F, top right), transgender women (G, bottom left), and non-binary people assigned male at birth (H, bottom right). Each colour corresponds to a different site, as indicated in the legend. Non-melanoma skin cancers are not included in this plot.

Expected cancer cases by anatomical site for all TGD people is shown in Figure 1B. The top 5 cancers expected are breast (19%, n = 187, 95% UI 149-241), colorectal (12%, n = 117, 95% UI 106-129), lung (11%, n = 108, 95% UI 96-122), melanoma (7.1%, n = 69, 95% UI 64-74) and urinary (6.2%, n = 60, 95% UI 54-67). Expected cases by anatomical site are shown for the four gender identity groups in Figure 2E-H. Breast, colorectal and lung cancer are expected to be in the top 5 most common cancer types for all four groups, though the order varied by gender. For comparison, the top 5 most common cancers in 2023 observed for cisgender persons were prostate (17%), breast (15%), lung (13%), colorectal (12%) and urinary (7%) (Supplementary Figure 3).

We next focused on three sex-specific cancer subtypes, breast, prostate and gynaecological, as GAC may affect cancer incidence. Therefore, we report expected cases based on the current access to GAC TGD individuals reported in 2025 and next, based on their reported planned uptake.

### Breast cancer

Of the total predicted cases of breast cancer with the current level of GAC (n= 187, 95% UI 149-241), 110 (95% UI 92-133) are expected to occur in transgender men, 43 (95% UI 25-68) in transgender women, 29 (95% UI 24-35) in non-binary people assigned female at birth, and 5 (95% UI 3-9) in non-binary people assigned male at birth.

With the current level of GAC access, the peak age of cases is expected to be 50-54 years in the transgender men and non-binary people assigned female at birth, accessing male chest reconstruction with or without concomitant hormone therapy (Figure 3A). Among transgender women and non-binary people assigned male at birth accessing feminising hormone therapy the peak age at diagnosis is expected to occur at ages 60-64 (though cases were very low in this latter category: n = 6, 95% UI 3-11 in transgender women and n=0.6, 95% UI 0.2-1.3 in non-binary people assigned male at birth) (Figure 3C).

**Figure 3.**
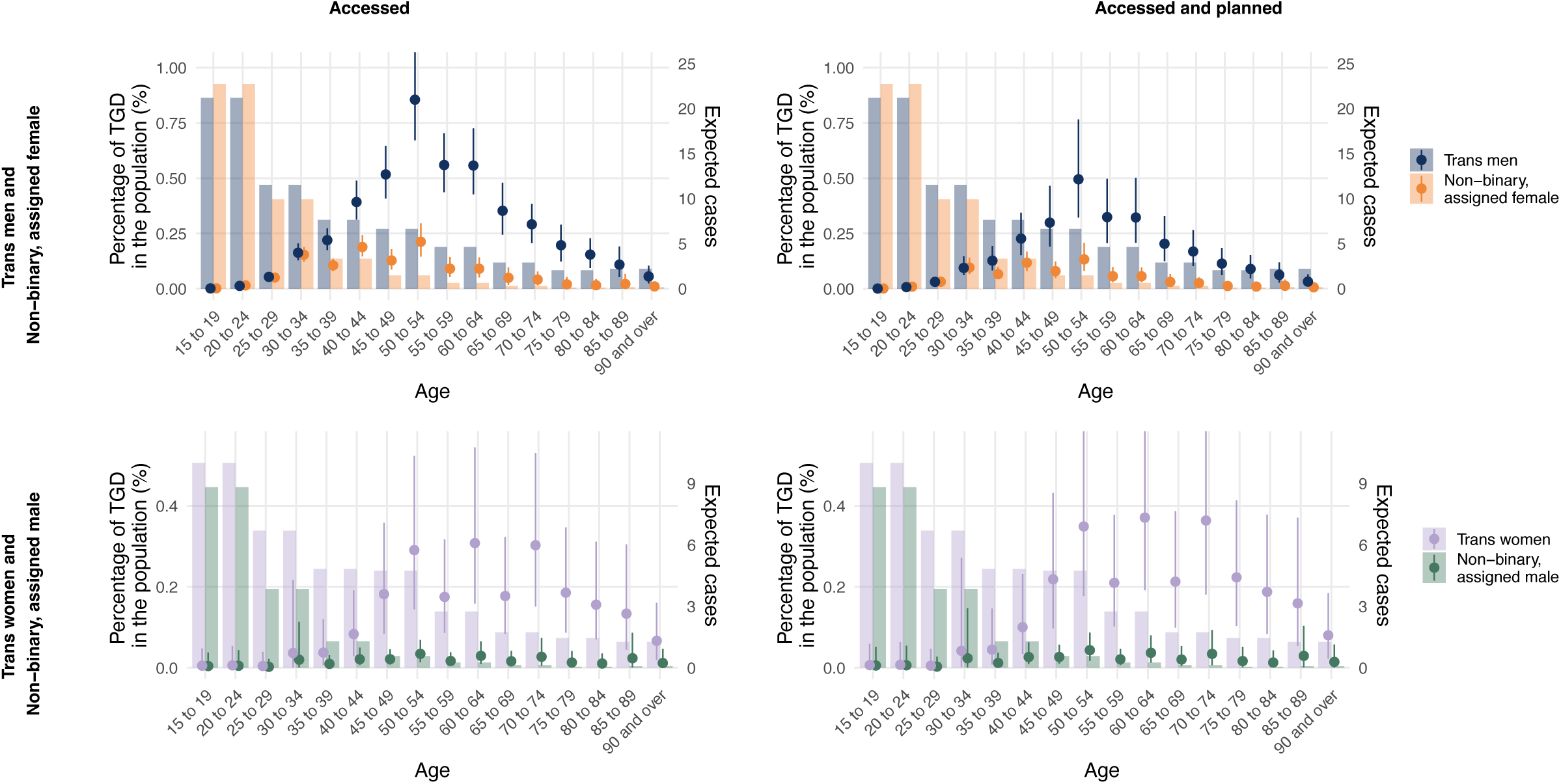
A-D. Proportion of TGD people in the population (left y-axis) and expected number of breast cancer cases (right y-axis) in transgender men (dark blue) and non-binary people assigned female at birth (orange), with current level of GAC uptake (A, top left) and planned level of uptake to GAC (B, top right); and in transgender women (dark green) and non-binary people assigned male at birth (purple), with current level of GAC uptake (C, bottom left) or planned level of uptake to GAC (D, bottom right). Bar plots show the age-stratified proportion of transgender men (dark blue), non-binary people assigned female at birth (orange), transgender women (dark green) and non-binary people assigned male at birth (purple) in the population, based on GPPS survey data. Each dot corresponds to the expected number of breast cancer cases in an age category (x-axis). Error bars show 95% uncertainty intervals for each age category.

When we estimate the number of cases based on planned GAC uptake, total breast cancer cases are expected to fall to 140 (95% UI 92-209). Cases are expected to reduce to 64 (95% UI 42-97) among transgender men and 18 (95% UI 13-26) among non-binary people assigned female at birth. Peak age of diagnosis is expected to remain at 50-54 years in both groups (Figure 3B). Cases are expected to increase to 51 (95% UI 29-82) in transgender women and 7 (95% UI 3-12) in non-binary people assigned male at birth, with peak age of diagnosis staying the same for both groups (Figure 3D).

### Prostate

Of the total predicted cases of prostate cancer with the current level of feminising hormone therapy access (n= 49, 95% UI 32-77), 44 (95% UI 28-68) were in trans women and 5 (95% UI 3-9) were in non-binary people who were assigned male at birth, with a highest number of cases expected to be in 60-74 year olds (Figure 4A).

**Figure 4.**
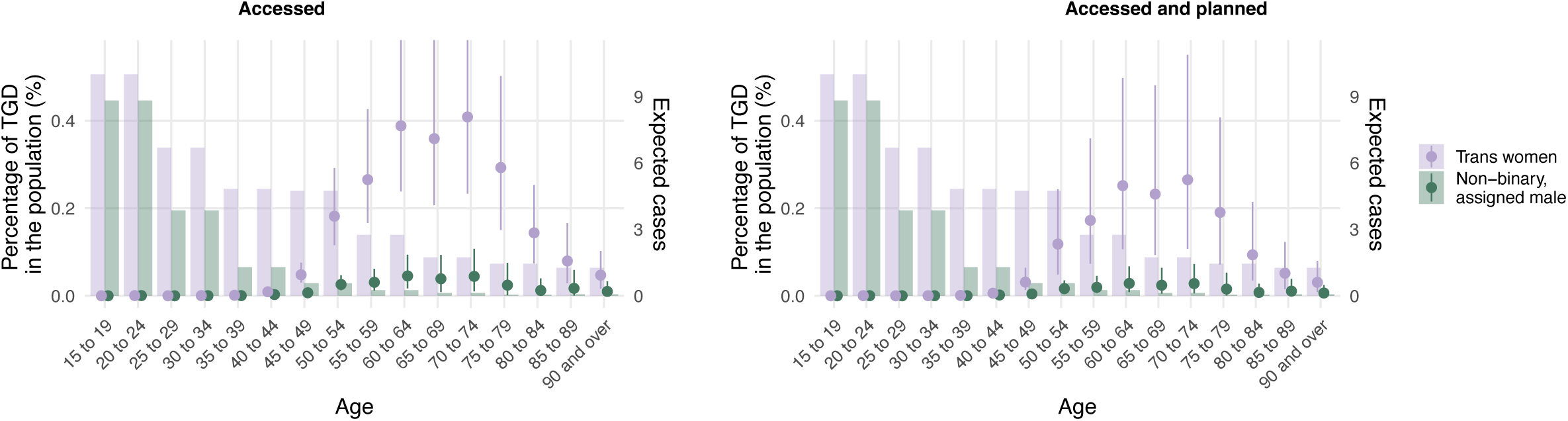
A-B. Proportion of TGD people in the population (left y-axis) and estimated number of prostate cancer cases in 2025 (right y-axis) in transgender women (dark green) and non-binary people assigned male at birth (purple), with current level of GAC uptake (A, left) or planned level of access to GAC (B, right). Bar plots show the age-stratified proportion of transgender women (dark green) and non-binary people assigned male at birth (purple) in the population, based on GPPS survey data. Each dot corresponds to the estimated number of breast cancer cases in an age category (x-axis). Error bars show 95% uncertainty intervals for each age category.

When we estimate the number of cases based on planned level of access to feminising hormone therapy, total prostate cancer cases are predicted to fall to n=32 (95% UI 14-64), 29 (95% UI 12-58) for trans women and 3 (95% UI 1-7) for non-binary people with peak age of diagnosis remaining at 70-74 years (Figure 4B).

### Gynaecological

We considered the three most common gynaecological cancers in cisgender women; uterus, ovary, and cervix.

At the current level of access to gender-affirming hysterectomy and bilateral salpingoophorectomy, expected total cases for uterine cancer in trans men and non-binary people assigned female at birth are 30 (95% UI 27-33) with an expected peak age of diagnosis of 60-64 in trans men and 50-54 in non-binary people (Figure 5A). When we estimate case numbers based on planned gender–affirming hysterectomy, the total expected cases falls to 7 (95% UI 5-9) across both groups with disappearance of a clear peak age due to low numbers (Figure 5B).

**Figure 5.**
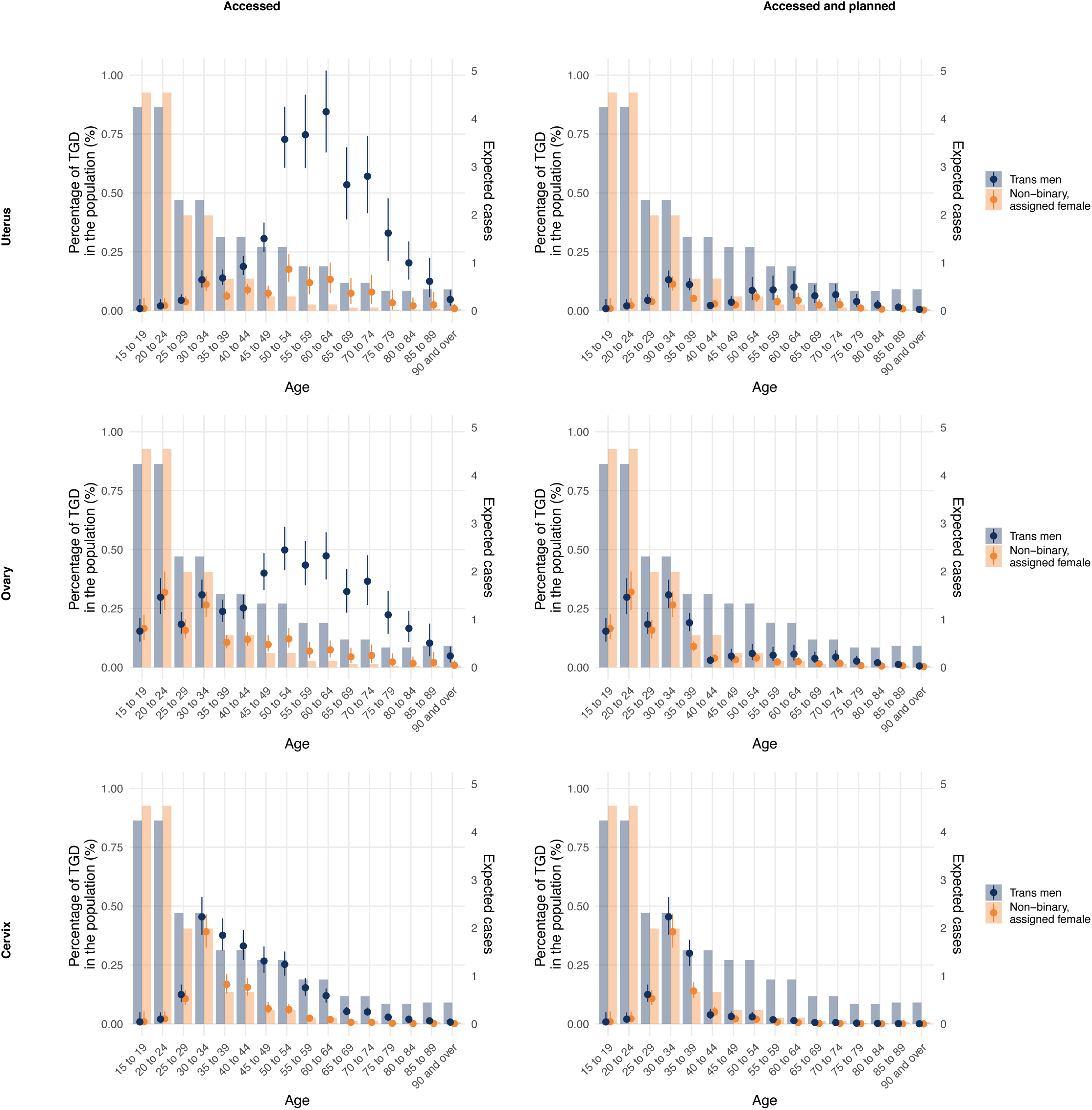
A-B. Proportion of TGD people in the population (left y-axis) and estimated number of uterus cancer cases in 2025 (right y-axis) in transgender men (dark blue) and non-binary people assigned female at birth (orange), with current level of GAC uptake (A, top left) or planned level of access to GAC (B, top right). C-D. Proportion of TGD people in the population (left y-axis) and estimated number of ovary cancer cases in 2025 (right y-axis) in transgender men (dark blue) and non-binary people assigned female at birth (orange), with current level of GAC uptake (A, top left) or planned level of access to GAC (B, top right). E-F. Proportion of TGD people in the population (left y-axis) and estimated number of cervix cancer cases in 2025 (right y-axis) in transgender men (dark blue) and non-binary people assigned female at birth (orange), with current level of GAC uptake (A, top left) or planned level of access to GAC (B, top right). For all plots: Bar plots (left y-axis) show the age-stratified proportion of transgender men (dark blue) and non-binary people assigned female at birth (orange) in the population, based on GPPS survey data. Each dot corresponds to the estimated number of breast cancer cases in an age category (x-axis). Error bars show 95% uncertainty intervals for each age category.

For ovarian cancer, total cases in trans men and non-binary people assigned female at birth are expected to be 30 (95% UI 28-32) with current access to hysterectomy with bilateral salpingoophorectomy, falling to 13 (95% UI 12-15) when we consider TGD individuals’ planned uptake. Peak age of cases is expected to be 50-64 years in trans men with current access and shifted to 20-34 years with planned uptake (Figure 5C, D).

Finally, with current access to hysterectomy, the total expected cases of cervical cancer across transgender men and non-binary people assigned female at birth will be 16 (95% UI 15-18). This decreased to 9 (95% UI 8-10) cases when considering planned uptake of hysterectomy. Under both assumptions, the peak age of diagnosis is predicted to be 30-34 years across both groups (Figure 5E, F).

### 5 and 10-year predictions

Including other cancers where GAC could affect risk for anatomical reasons (testicular, penile and other gynaecological cancers), if TGD people being able to access their planned level of GAC reported in 2025, this would reduce the total cancers (excluding non-melanoma skin) from 966 to 853 (95% UI 761-972).

Assuming the same level of GAC access as reported in 2025 and the principal census scenario, our predictions estimate the total cases of cancer in TGD people to be 1740 (95% UI 1584-1934) in 5 years and 2258 (95% UI 2066-2507) in 10 years (excluding non-melanoma skin). Improvements in availability of GAC to allow all TGD people to access their planned GAC at a similar age would reduce these totals to 1555 (95% CI 1386-1766) at 5 years and 2012 (95% CI 1797-2282) at 10 years (Figure 6, Supplementary table 6).

**Figure 6.**
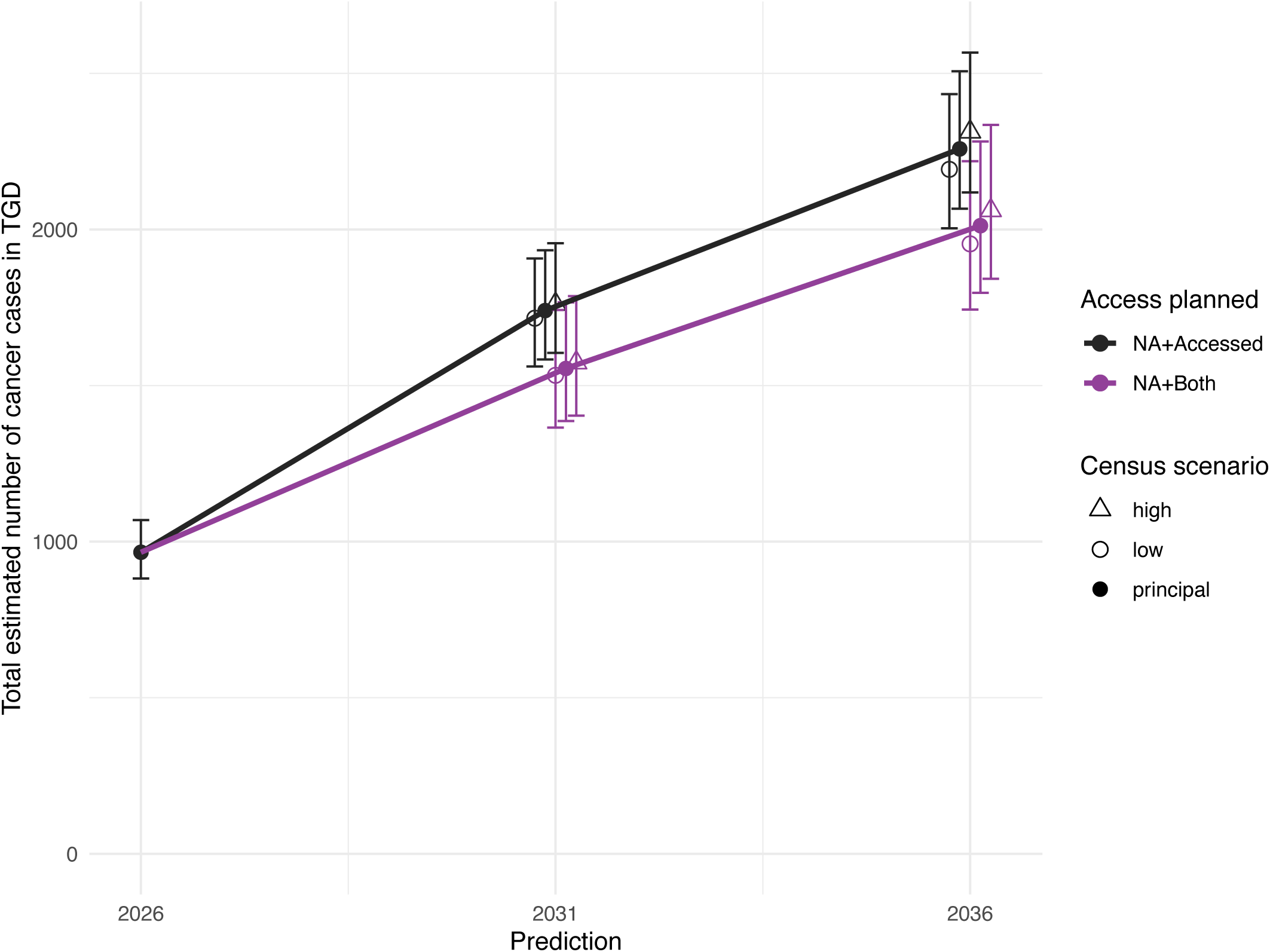
Estimated number of cancer cases in 2025 in TGD people and estimated predictions in 5 and 10 years, either with the same level of GAC access as in 2025 (black line), or with the planned GAC (purple line). Error bars show 95% uncertainty intervals. Symbols corresponds to three different census scenarios: principal, low fertility, mortality and migration and high fertility, mortality and migration.

## Discussion

We predict there are likely to be approximately 966 cases of cancer (excluding non-melanoma skin) in TGD people per year in England. Most cases are expected to occur in transgender men, followed by transgender women, and non-binary people. Case numbers are expected to be highest for breast, lung, and colorectal cancers, melanoma and lymphoma. This differs from the top five most common cancers in cisgender people likely owing to a younger age distribution, and the reduction in prostate cancer incidence owing to feminising hormone therapy with androgen deprivation.[10]

Access to GAC is predicted to reduce total cases of breast, prostate, gynaecological cancers, though there is likely to be a small increase in breast cancer cases in transgender women and non-binary people who were assigned male at birth.

Improved access to GAC to allow interventions currently desired or planned by TGD people but not yet undertaken could reduce total expected cases in 10 years from 2258 to 2012 (an 11% reduction). To our knowledge, this is the first study to attempt to predict the number of cancer cases for TGD people in any country. It is not currently possible to validate these predictions due to lack of recording of gender incongruence in cancer registries, but this makes such estimates all the more important to inform training and service provision. It also highlights the urgent need for sensitive collection of this data.

We base our population estimates on a combination of Census and GPPS data in order to overcome potential inaccuracies in self-reporting of gender incongruence.[28] We used a community survey to estimate current access and planned uptake of GAC because many TGD people access GAC privately or by self-medicating, meaning the survey is likely more accurate than data from primary care or gender identity services.

A major strength of our study is the use of Monte Carlo simulations, a stochastic method that accounts for uncertainty in data from multiple sources (e.g., cancer incidence, GAC rates, and TGD population size) and generates robust 95% uncertainty intervals, providing more reliable ranges. Furthermore, our method considers the impact of surgery by simulating an age-dependent risk (with SIR=0 post-median surgery age) and avoids the underestimation of rare events by using log-uniform distributions in cases where incidence = 0 with confidence intervals different from 0. Our predictions rely on a validated model for cancer incidence and multiple scenario analyses using census projections.

Our study has several limitations. Firstly, we base cancer case estimates for each TGD subgroup on the age-adjusted incidence for the cisgender group within the same sex assigned at birth. We also assume the incidence rate ratio is the same across all age strata (though, age of commencing hormones and length of hormone exposure will likely alter the age-adjusted incidence of cancer). These nuances are challenging to model with insufficient data as there are few cohort-based epidemiological studies of cancer incidence in TGD confirmed to have accessed hormone therapy, from which to adjust risks. We also had to assume a single SIR for individuals undergoing male chest reconstruction, regardless of testosterone use. Our predictions are based using current GPPS data and do not account for potential future changes in the prevalence of TGD populations. We also assume that all TGD people will disclose their gender incongruence in the GPPS which may not be the case, particularly for older populations who have experienced historical stigma and discrimination.

We also made several assumptions for the GPPS data, which reports 10 year age categories. We deduced the disclosure per 5-year category by dividing the numbers by 2. This assumes a uniform distribution of TGD people across each 10-year age category. For non-binary people and people who prefer to self-describe, sex assigned at birth is not available in GPPS.[1] We therefore applied the proportions of people identifying as non-binary for each sex assigned at birth from the census. This does not account for potential age variations among these groups. The community survey on GAC was a convenience sample, it might not be representative of access among TGD individuals in the UK, though it achieved good geographical distribution. Our estimation of cases at 5 and 10 years makes several assumptions. These include that disclosure of TGD identity for age groups will remain constant as they age, that the youngest age categories will have the same level of disclosure as those categories do in 2025 and that desires for GAC and median age of access will remain constant.

Finally, we were unable to adjust for other cancer risk factors, such as smoking and HIV infection, despite the fact that they are known to be higher in some subgroups of TGD people in the UK.[8] This also means that our 5- and 10-year predictions do not adjust for changes in health behaviours. For example, gender-neutral HPV vaccination will likely result in a reduction in cases of cervical and other HPV-related cancers for genders. Though no other studies have attempted to estimate cancer cases in TGD people, several epidemiological studies have sought to estimate incidence, including those used to derive the SIRs used in this study. We were also unable to adjust for other demographics and social determinants of health. For example, lower socioeconomic status is linked to cancer risk and TGD people are known to be 2.5 times were likely to be in the lowest socioeconomic group.[29]

Researchers at Amsterdam UMC have carried out several longitudinal studies linking health records from their national gender clinic to their nationwide pathology database, restricting this to those accessing GAC.[9–12,24,30] They found a higher SIR for breast cancer and meningioma in transgender women on feminising hormone therapy, a lower SIR for breast cancer in transgender men on masculinising hormone therapy with or without male chest reconstruction, and a lower SIR for prostate cancer among transgender women accessing feminising hormone therapy.[9,10,24] They found no difference in testicular or gynaecological cancer incidence.[11,12,30] However, sterilisation was mandatory for legal gender change in the Netherlands until 2014[31] which limits the ability to study these cancer types among TGD people with differing access to GAC.

A study conducted amongst US Veterans found no added risk of breast cancer for transgender women, with and without hormone therapy compared to cisgender men in the US population.[32] They also found no reduced breast cancer risk for transgender men compared to cisgender women, contrary to the results of the Dutch study, likely due in part to the substantially lower follow-up time in the US. Additional data from the US Veterans Affairs suggest that prostate cancer incidence is lower among transgender women than for cisgender men, in line with the Dutch results.[33]

These studies were conducted in countries with differing GAC protocols and access from the UK, making it difficult to draw conclusions about expected cancer incidence and case numbers in England. While our estimates provide a reliable indicator, the gold standard would be recording of gender incongruence in NCRAS,[34] necessitating sensitive and consistent data collection in secondary care. However, case estimation alone allows planning for specialist and integrated care provision.

Cancer in TGD people is often regarded as a rare occurrence as they are a minoritised population. However, at an estimated 966 cases per year England in 2025, this is far more common than primary cancers in pregnancy, where 84 cases were identified over a two-year period 2015-2017 in the entire UK[35] or even (presumed cisgender) male breast cancer, with 341 cases in England in 2019.[22] Yet these two scenarios appear more commonly in oncology professional training curricula and conference programmes. A 2020 survey of UK oncologists found that only 57% thought that it was important to know a patient’s gender identity and only 3% regularly asked and the same has been found internationally. [36–39] It is vital that all healthcare professionals receive appropriate training not only around sensitive communication with transgender and gender diverse people (e.g. name and pronoun use,[40] how to talk about sex-specific body parts and discuss needs and preferences around gender-affirming care) but also how this gender-affirming care might specifically intersect with a cancer diagnosis.[41,42] Such education has already been recommended in a joint statement led by the Royal College of Radiologists.[43]

In oncology, TGD people also have additional medical needs including balancing the safety of gender-affirming hormone therapies when diagnosed with hormone-responsive cancers,[44–46] planning of radiotherapy,[47] and dosing and monitoring of systemic anti-cancer therapies.[46,48] They also experience additional support needs, including concerns around embodiment after cancer, psychosexual wellbeing and end-of life.[49–51]

Breast cancer is a key example where treatment has multiple intersections with GAC. For trans men and non-binary people who were assigned female at birth, breast tumours may be found incidentally at bilateral male chest reconstruction,[45] interrupting gender transition while staging and treatment are undertaken.[52] Testosterone can be metabolized into oestrogen, necessitating adjustments to endocrine therapy in oestrogen receptor (ER) positive tumours. In androgen receptor positive tumours, testosterone appears to have potentially tumour-suppressive effects in ER-positive tumours and growth promoting effects in ER-negative tumours. In transgender women and non-binary people who were assigned male at birth who have breasts, mastectomy may cause gender dysphoria and patients may be advised to discontinue feminizing hormones if their tumour is ER-positive. With a predicted 187 cases of breast cancer in TGD people per year it appears that healthcare professionals in breast oncology are a priority for training.

Cancers in non-sex specific organs may also be of special consideration in trans people. For example, in colorectal cancer, previous cancer surgeries or radiotherapy may impact future genital surgeries.[46] In haematological cancers, hormone therapy may need to be adjusted due to bleeding or thrombosis risk.[46] Expertise in these intersections is currently being amassed by the UK Cancer and Transition Service, a national hybrid clinic and multidisciplinary team meeting for TGD people with cancer in the UK that facilitates shared decision making with patients, while upskilling local teams and providing a platform for data collection.

As well as facilitating improved research and training around management of cancer for TGD individuals, understanding what types of cancer they experience allows us to focus on preventative efforts. While this paper did not adjust for cancer risk and preventative factors, emphasis on smoking cessation, healthy diets, HPV vaccination, and avoidance of UV exposure would be worthwhile campaigns in TGD communities given that lung, colorectal and melanoma skin cancers predicted to be the 2^nd^, 3^rd^ and 4^th^ most common cancer types for this population. With breast being the most common cancer, and its risk being modulated by hormone therapy, age-adjusted incidence data would help determine if adjustments are needed to national screening for TGD people accessing hormones, beyond simply accounting for whether they have breasts. As no study to date has been large enough to do this, it should be a focus for future research.

Lastly, while reduction in cancer risk should not be a primary driver in gender-affirming surgeries[5] because of any increased cancer risk, it is a factor considered by many TGD people in their decision making, given potential gender dysphoria resulting from a cancer diagnosis and treatment, as well as any recommended screenings. This study shows that timely access to the GAC desired by gender diverse people in the UK is likely to substantially reduce the total number of cases of cancer, simply due to changes in anatomy and hormonal milieu.

Increased GAC provision and efficiency within the NHS, as called for by the review of adult gender services in England,[14] is also likely to improve emotional wellbeing in the TGD population.[53,54] This has the potential to reduce cancer through lessening of improving uptake of health-promoting behaviours and preventative care.[55,56] This not only benefits the individual but is a cost-saving to the NHS.

This study is not a substitute for appropriate recording of sex characteristics, gender identity and gender incongruence in healthcare system databases. These data are necessary to derive high-quality incidence data for TGD people, and enable oncologists to tailor care to individual patients, but also to identify discrepancies that point to underlying inequities.

Collection of these data must be accompanied by appropriate training for healthcare and administrative staff, appropriate functionality in the electronic health record and community engagement to ensure that these data are collected in a way that is acceptable with appropriate legal and ethical safeguards that will build trust. The importance to the individual, and benefit to the TGD community, also need to be communicated though work with charity and community organisations.

### Conclusion

This study provides prediction of cancer cases in TGD people in England, supporting the planning of service provision and training. This is vital, as with increasing disclosure, and long wait times for GAC, cancer cases in TGD people are predicted to increase. Our findings demonstrate a net-reduction in cancer cases in TGD people if they are able to access the specific gender-affirming medical interventions they have planned. This makes timely access to GAC both potentially life-saving to the individual and cost-saving to the NHS.

## Acknowledgements

We would like to thank respondents to the community health survey for their participation, without which we would not have been able to obtain informed estimates of gender affirming care use.

## Authors’ contributions

AMB and SJ defined the study concept. TW extracted and cleaned data from NCRAS. AMB and CP extracted and cleaned data from GPPS and Census. L-ET and BM conducted the community survey and extracted data for use in this study. CP carried out the statistical analysis and created the figures. CP, AMB and SJ drafted the manuscript. All authors reviewed and refined the manuscript. The corresponding author attests that all listed authors meet authorship criteria and that no others meeting the criteria have been omitted. AMB is the guarantor.

## Statement

AMB has the right to grant on behalf of all authors and does grant on behalf of all authors, an exclusive licence (or non exclusive for government employees) on a worldwide basis to the BMJ Publishing Group Ltd to permit this article (if accepted) to be published in BMJ editions and any other BMJPGL products and sublicences such use and exploit all subsidiary rights, as set out in our licence.

## Transparency declaration

AMB affirms that the manuscript is an honest, accurate, and transparent account of the study being reported; that no important aspects of the study have been omitted; and that any discrepancies from the study as planned (and, if relevant, registered) have been explained.

## Ethics approval and consent to participate

Not applicable.

## Patient and Public Involvement

This study was conceived through feedback from patient and public involvement focus groups with TGD patients with cancer. These were carried out with facilitation of the OUTpatients LGBTIQ+ Cancer Charity. The OUTpatients CEO has lived experience as a non-binary person with cancer and is a co-author on this paper. Multiple authors on this paper are also part of the TGD community.

## Data availability

We used publicly available datasets. They are available alongside our R codes: https://github.com/chlpasin/Estimation-of-cancer-cases-in-TGD-people-in-England.

Further Summary data for the survey on access to GAC will be published in a subsequent dedicated manuscript but raw data from individual respondents will not be made available to maintain anonymity.

