## Supplemental materials for "Estimation of cancer cases in transgender and gender diverse people in England"

**Supplementary methods**

Data sources

*Age- and sex-specific denominators for cisgender people*

Those were obtained from the Census 2021 counts through the Office of National Statistics (ONS). To obtain the denominator for the age-specific proportion of TGD people in England, we used the NHS GP Patient Survey (GPPS) 2025. We chose not to use data from the ONS for age-specific statistics on TGD people for two reasons.

Firstly, there was significant controversy over how questions on sex and gender were asked. Guidance for the question “What is your sex?” initially instructed individuals to use the sex marker on their passport. This was changed following a court order due to “If you are considering how to answer, use the sex recorded on your birth certificate or Gender Recognition Certificate,” because passports can be changed without legal process.[1] A follow-up question on gender asked people “Is the gender you identify with the same as your sex registered at birth?”, with the option of selecting either “Yes”, or “No” and writing in their gender identity. As a result of these changes, some trans people threatened to boycott the Census 2021 altogether, or the sex and/or gender questions, due to both privacy concerns, lack of ability to self-identify their sex as per their passport, and the lack of clarity in the question (if purpose of the Census 2021 sex question was to capture the sex registered at birth, this will not be correct if someone uses the sex on their gender recognition certificate).[2,3] Six percent of the population declined to answer the question on gender in the ONS Census 2021, compared to only 0.87% in GPPS 2025.

Secondly, following publication of the ONS Census data, there have been concerns that the question was misinterpreted by those who did not speak English as a first language.[4]

In the GPPS data, we classified people responding “male” (respectively “female”) to the question “which of the following best describes you?” and “No” to the question “is your gender identity the same as the sex you were registered at birth” as “trans men” (respectively “trans women”). People responding “non-binary” or “prefer to self-describe” were classified as “non-binary and gender non-conforming”. Non-binary people were further divided in two categories, assigned male or female at birth. Based on estimates from the ONS,[5] we assigned 67.5% of non-binary people as female at birth 32.7% of as male at birth.

*Access to gender-affirming care*

We relied on an online mixed-methods community survey. This was a separate project conducted by authors L-E T and BM and aimed to better understand the experiences of transgender, non-binary and gender-diverse people over the age of 18 seeking access to, currently accessing or having accessed gender-affirming healthcare in England.The project was approved by the Lancaster University Medical School Postgraduate Taught Director, as delegated by the Lancaster University Medical School Faculty Ethics Committee (Case ID: MCR2425EA02THYNNE). Informed consent was provided by all participants. Participants were recruited opportunistically through social media and promoted by non-profit organisations. The survey was open between 19^th^ January – 6^th^ April 2025, with 419 responses eligible for final analysis.

The number of people who accessed, wanted to access and did not want to access hormone replacement therapy and gender-affirming surgeries was determined by the question “What types of gender-affirming healthcare are you seeking?” Gender identities were identified using the checklist question “What gender or which genders do you currently identify with?”. If a participant identifies as non-binary, their answer to the question “Are you seeking/have you sought predominantly masculinising or feminising gender-affirming care?” was used to identify the healthcare interventions they may wish to access. Age was collected using age brackets.

Number of participants who had already access and total number had access and planned to access relevant interventions was calculated across four identity categories (trans men, trans women, non-binary people assigned female at birth and non-binary people assigned male at birth). Median age for accessing each intervention was calculated for each intervention within each identity category using the mid-point of the prescribed age bracket for each respondent.

**Supplementary Figure 1**

A-D. Histogram showing the estimated TGD population by age category in 2025 (y-axis) in by gender identity and sex assigned at birth; trans men (A, top left, blue), non-binary people who were assigned female at birth (B, top right, orange), trans women (C, bottom left, green), non-binary people who were assigned male at birth (D, bottom right, purple). Each bar corresponds to an age category (x-axis). Error bars show 95% uncertainty intervals for each age category.

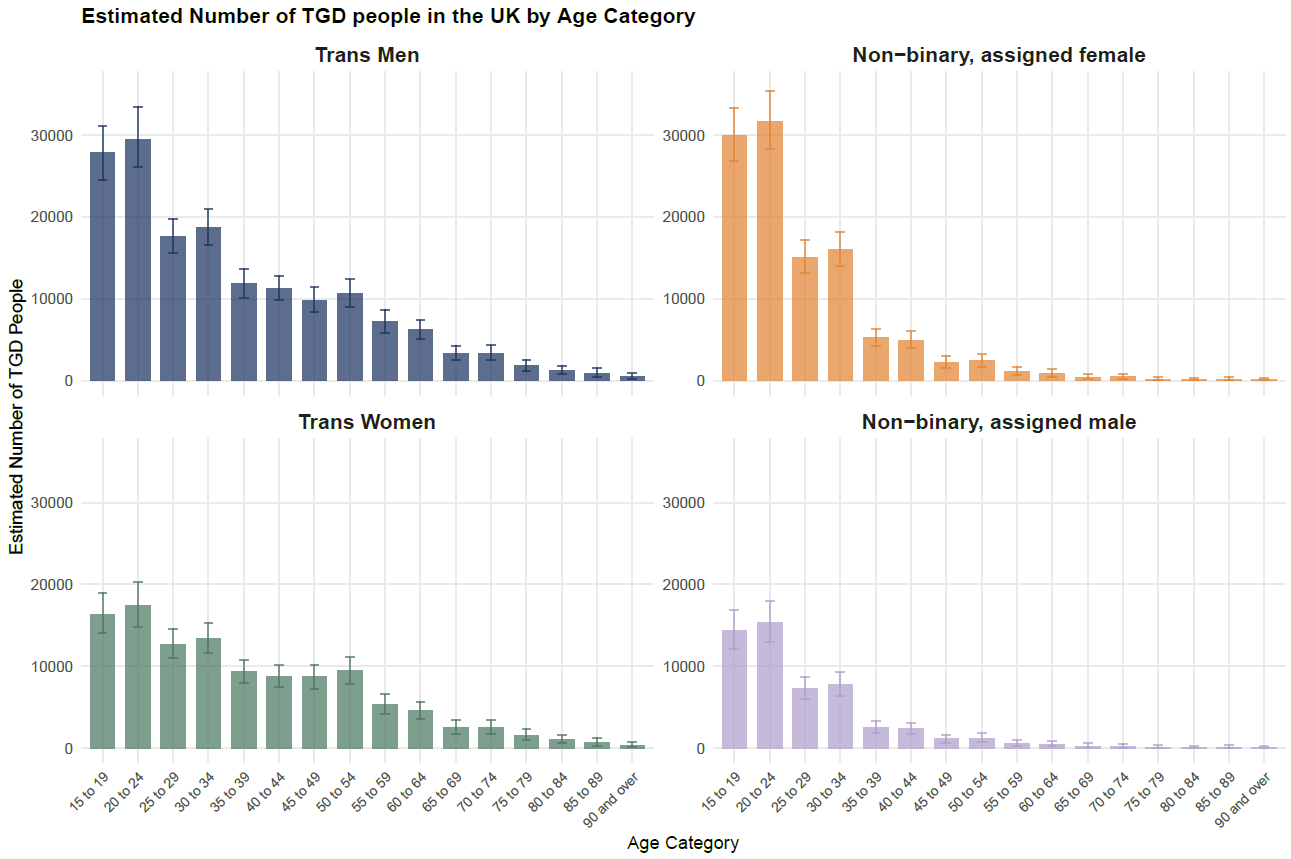

**Supplementary Figure 2**

A-C. Histogram showing the estimated number of cancer cases in 2025 (y-axis) in cisgender female (A, top left, purple), cisgender male (B, top right, green), all cisgender (C, bottom). Each bar corresponds to an age category (x-axis). Error bars show 95% uncertainty intervals for each age category. Dark color shows the number of cases across all cancer type, except non-melanoma skin cancer. Light color shows the total number of cases (including non-melanoma skin cancer).

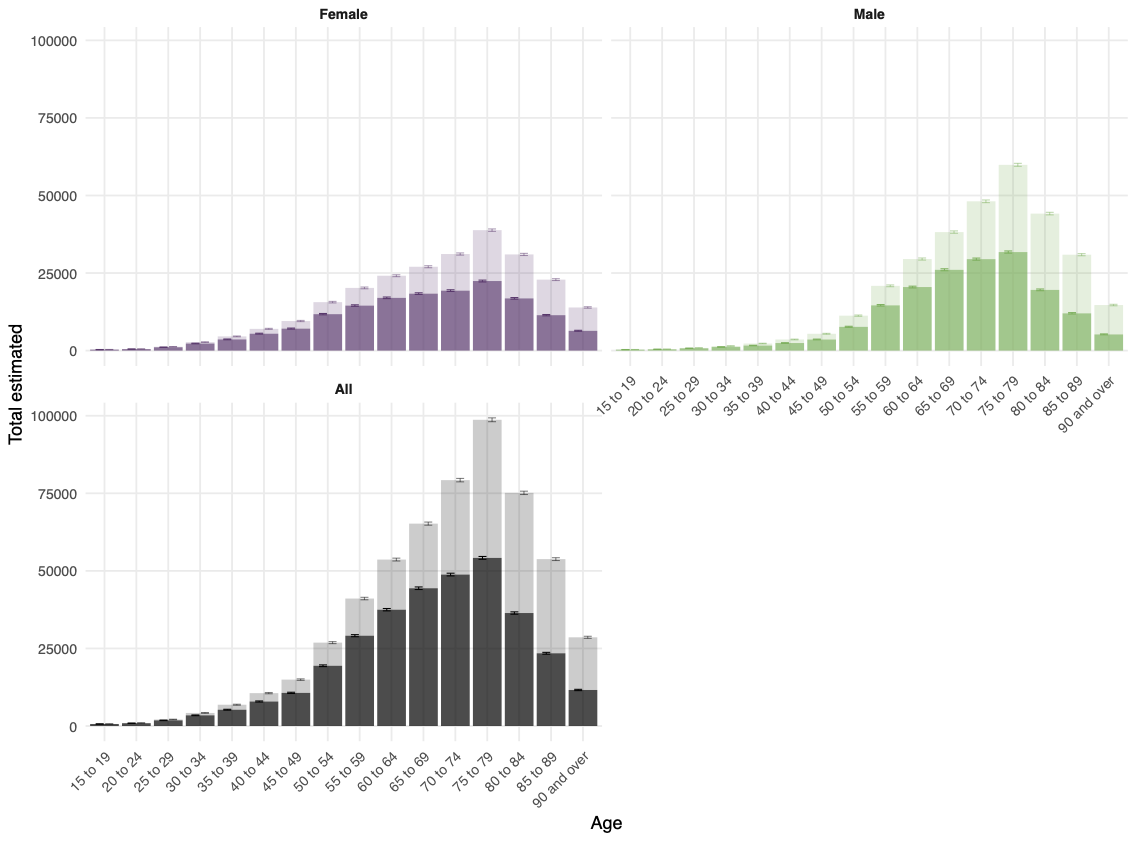

**Supplementary Figure 3**

Pie chart depicting the proportion of total estimated number of cancer cases by anatomical site in all cisgender people. Each color corresponds to a different site, as indicated in the legend. Non-melanoma skin cancers are not included in this plot.

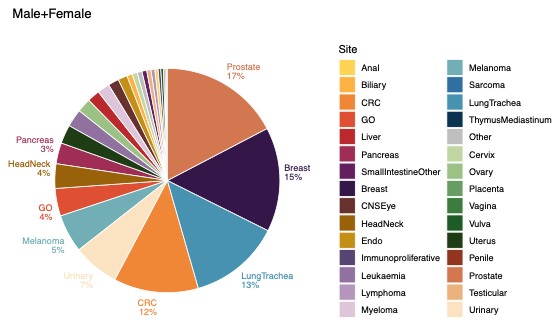
