## Supplementary Tables 1-3 for "Estimation of cancer cases in transgender and gender diverse people in England"

**Supplementary Table 1 - ICD-10 Codes for Primary Malignant Tumours and Tumour Type Anatomical Groupings**

| **Tumour Type** | **ICD-10 Code** | **Site Description** |
| --- | --- | --- |
| **Head and Neck** | C00 C01 C02 C03 C04 C05 C06 C07 C08 C09 C10 C11 C12 C13 C14 C30 C31 C32 | Malignant neoplasm of lip Malignant neoplasm of base of tongue Malignant neoplasm of other and unspecified parts of tongue Malignant neoplasm of gum Malignant neoplasm of floor of mouth Malignant neoplasm of palate Malignant neoplasm of other and unspecified parts of mouth Malignant neoplasm of parotid gland Malignant neoplasm of other and unspecified major salivary glands Malignant neoplasm of tonsil Malignant neoplasm of oropharynx Malignant neoplasm of nasopharynx Malignant neoplasm of pyriform sinus Malignant neoplasm of hypopharynx Malignant neoplasm of other and ill-defined sites in lip, oral cavity and pharynx Malignant neoplasm of nasal cavity and middle ear Malignant neoplasm of accessory sinuses Malignant neoplasm of larynx |
| **Gastro-oesophageal** | C15 C16 | Malignant neoplasm of oesophagus Malignant neoplasm of stomach |
| **Small Intestine/Other** | C17 C26 | Malignant neoplasm of small intestine Malignant neoplasm of other and ill-defined digestive organs |
| **Colorectal** | C18 C19 C20 | Malignant neoplasm of colon Malignant neoplasm of rectosigmoid junction Malignant neoplasm of rectum |
| **Anal** | C21 | Malignant neoplasm of anus and anal canal |
| **Liver** | C22 | Malignant neoplasm of liver and intrahepatic bile ducts |
| **Biliary** | C23 C24 | Malignant neoplasm of gallbladder Malignant neoplasm of other and unspecified parts of biliary tract |
| **Pancreas** | C25 | Malignant neoplasm of pancreas |
| **Thymus/Mediastinum** | C37 C38 C39 | Malignant neoplasm of thymus Malignant neoplasm of heart, mediastinum and pleura Malignant neoplasm of other and ill-defined sites in respiratory system and intrathoracic organs |
| **Sarcoma** | C40 C41 | Malignant neoplasm of bone and articular cartilage of limbs Malignant neoplasm of bone and articular cartilage of other and unspecified sites |
| **Lung/Trachea** | C33 C34 C45 | Malignant neoplasm of trachea Malignant neoplasm of bronchus and lung Mesothelioma |
| **Melanoma** | C43 | Malignant melanoma of skin |
| **Non-Melanoma Skin** | C44 C46 | Other and unspecified malignant neoplasm of skin Kaposi sarcoma |
| **Breast** | C50 | Malignant neoplasm of breast |
| **Vulva** | C51 | Malignant neoplasm of vulva |
| **Vagina** | C52 | Malignant neoplasm of vagina |
| **Cervix** | C53 | Malignant neoplasm of cervix uteri |
| **Uterus** | C54 C55 | Malignant neoplasm of corpus uteri Malignant neoplasm of uterus, part unspecified |
| **Ovary** | C56 C57 | Malignant neoplasm of ovary Malignant neoplasm of other and unspecified female genital organs |
| **Placenta** | C58 | Malignant neoplasm of placenta |
| **Penile** | C60 | Malignant neoplasm of penis |
| **Prostate** | C61 | Malignant neoplasm of prostate |
| **Testicular** | C62 C63 | Malignant neoplasm of testis Malignant neoplasm of other and unspecified male genital organs |
| **Urinary** | C64 C65 C66 C67 C68 | Malignant neoplasm of kidney, except renal pelvis Malignant neoplasm of renal pelvis Malignant neoplasm of ureter Malignant neoplasm of bladder Malignant neoplasm of other and unspecified urinary organs |
| **CNS/Eye** | C69 C71 C72 | Malignant neoplasm of eye and adnexa Malignant neoplasm of brain Malignant neoplasm of spinal cord, cranial nerves and other parts of central nervous system |
| **Endocrine** | C73 C74 C75 | Malignant neoplasm of thyroid gland Malignant neoplasm of adrenal gland Malignant neoplasm of other endocrine glands and related structures |
| **Lymphoma** | C81 C82 C83 C84 C85 C86 C96 | Hodgkin lymphoma Follicular lymphoma Non-follicular lymphoma Mature T/NK-cell lymphomas Other and unspecified types of non-Hodgkin lymphoma Other specified types of T/NK-cell lymphoma Other and unspecified malignant neoplasms of lymphoid, haematopoietic and related tissue |
| **Immunoproliferative** | C88 | Malignant immunoproliferative diseases and certain other B-cell lymphomas |
| **Myeloma** | C90 | Multiple myeloma and malignant plasma cell neoplasms |
| **Leukaemia** | C91 C92 C93 C94 C95 | Lymphoid leukaemia Myeloid leukaemia Monocytic leukaemia Other leukaemias of specified cell type Leukaemia of unspecified cell type |
| **Other** | C47 C48 C49 C76 C80 C97 | Malignant neoplasm of peripheral nerves and autonomic nervous system Malignant neoplasm of retroperitoneum and peritoneum Malignant neoplasm of other connective and soft tissue Malignant neoplasm of other and ill-defined sites Malignant neoplasm without specification of site Malignant neoplasms of independent (primary) multiple sites |

**Supplementary Table 2 – Gender affirming care interventions access and planned, according to a community health survey.** Note, testosterone therapy information is not supplied as it was not directly relevant to the risk calculations given the subtotal mastectomy as part of male chest reconstruction is likely to play a greater role in risk modulation.

| Intervention | Patient Group | Accessed (%) | Planning to Access (%) | Median Age of Access (years) |
| --- | --- | --- | --- | --- |
| *Male chest reconstruction* | Trans men | 46 | 35 | 28 |
|  | Non-binary people assigned female at birth | 38 | 34 |  |
| *Hysterectomy and bilateral salpingoophorectomy* | Trans men | 10 | 80 | 38 |
|  | Non-binary people assigned female at birth | 3 | 66 |  |
| *Masculinising genital surgery (phalloplasty or metoidioplasty)* | Trans men | 3 | 53 | 38 |
|  | Non-binary people assigned female at birth | 2 | 38 |  |
| *Femising hormone therapy (oestradiol and/or androgen blockade)* | Trans women | 83 | 17 | 33 |
|  | Non-binary people assigned male at birth | 78 | 20 |  |
| *Orchiectomy without feminising genital surgery* | Trans women | 16 | 2 | 43 |
|  | Non-binary people assigned male at birth | 6 | 43 |  |
| *Feminising genital reconstructive surgery (vaginoplasty/vulvoplasty)* | Trans women | 18 | 76 | 43 |
|  | Non-binary people assigned male at birth | 6 | 59 |  |

Supplementary Table 3 – Post-gender affirming care standardised-incidence ratio obtained from the literature.

| Tumour-type  (reference) | Patient Groups | Reference Group | Intervention | SIR (95% CI) |
| --- | --- | --- | --- | --- |
| *Breast*  *(de Blok et al,* 2019) | Trans men and non-binary people who were assigned female at birth | Cis women | Testosterone +/- male chest reconstruction* | 0.2  (0.1 to 0.5) |
|  | Trans women and non-binary people who were assigned male at birth | Cis men | Oestradiol +/- antiandrogen | 46.7  (27.2 to 75.4) |
| *Prostate*  *(de Nie et al, 2020)* | Trans women and non-binary people who were assigned male at birth | Cis men | Oestradiol +/- antiandrogen | 0.2  (0.08-0.42) |

* Given gender affirming care practices in Amsterdam, it is assumed that the majority will have undergone mastectomy and that this confers a large proportion of the breast cancer risk reduction.
